# Breaking the Cycle: Addressing the Drivers of Impoverishing Healthcare Costs in Georgia

**DOI:** 10.1101/2025.01.31.25321173

**Authors:** Tsotne Gorgodze, Akaki Zoidze, Jolly Mae Catalan, George Gotsadze

## Abstract

**Background:** Financial protection is a core goal of universal health coverage, ensuring access to necessary healthcare without financial hardship. It is assessed through indicators like catastrophic and impoverishing out-of-pocket (OOP) health spending. Despite Georgia’s Universal Health Coverage Program (UHCP) introduced in 2013, covering nearly 90% of the population, the incidence of catastrophic and impoverishing health spending remains high and stagnant compared to other European countries.

**Objective:** This study investigates factors associated with impoverishing health expenditures among Georgian households to inform policy decisions and prevent healthcare-related poverty.

**Methods:** Data from the Household Income and Expenditure Surveys (HIES) conducted by GeoStat from 2009 to 2023 (N = 198,292 households) were analyzed using survey-weighted logistic regression to account for the complex survey design. Impoverishing health expenditure, based on the relative poverty line suggested by the WHO Regional Office for Europe, was the outcome variable, with explanatory variables guided by Andersen’s behavioral model for healthcare utilization.

**Results:** OOP expenditure on drugs emerged as the most significant determinant of impoverishment (OR 43.3, 95% CI 39.3-47.7, P < 0.001). The poorest quintile faced the highest risk (OR 44.5, 95% CI 22.1-89.7, P < 0.001) compared to the richest quintile, with slightly lower odds for the second quintile. Impoverishment probabilities decreased before UHCP’s introduction (2009–2013) and slightly increased afterward, averaging 0.34 for 2014-2023 compared to 0.28 in 2013.

**Discussion:** The decline in impoverishment odds during 2009-2013 highlights the greater financial protection provided by benefits focused on the poor before UHCP’s expansion. Addressing impoverishing expenditures, particularly for outpatient drugs benefiting the poorest, requires increased public investment, broader coverage, and gradually extending benefits to other vulnerable groups. Incorporating lessons from the WHO Regional Office for Europe’s recommendations could further strengthen Georgia’s financial protection policies.

**Key Messages:** *What is already known on this topic:* After the introduction of UHCP in Georgia and many lower-middle-income countries, the progress towards better financial protection has been limited, undermining the success in service coverage index on the path to universal health coverage.

*What this study adds:* This study examines the factors dragging financial protection behind by exploring the drivers of healthcare costs, particularly types of health spending leading to health disparities.

*How this study might affect research, practice or policy:* This study provides policymakers with insights into drivers of impoverishing health expenditure, particularly how to curb OOPs on pharmaceuticals, especially for poor and reduce financial burden leading to impoverishment. The study suggests policy options for reducing impoverishing effects of spending on medicines and advocates for “progressive universalism”.

## Introduction

Financial protection, alongside service coverage, is a key objective of Universal Health Coverage (UHC) and is crucial for enhancing health and well-being for everyone. It also plays a vital role in advancing the broader goals of the 2030 Agenda for Sustainable Development and supports the commitment to leaving no one behind (1). These priorities align with the three fundamental dimensions of a country’s progress toward achieving UHC: population coverage, service coverage, and the proportion of healthcare costs covered. Advancing in these areas aims to reduce reliance on out-of-pocket (OOP) payments as a dominant source of healthcare funding (2). Even for nations often seen as having achieved UHC objectives, navigating trade-offs between the three UHC dimensions is necessary, making the future policy changes context-specific (3).

While globally service coverage improvements have been notable in previous years, progress in financial protection has not kept pace. Since 2000, nearly all countries have experienced improvements in service coverage, whereas catastrophic spending either worsened or showed minimal change in most countries (1), including Georgia (4). Therefore, this research examines the factors determining the impoverishing effects of OOPs in Georgia and provides ideas on how these insights can guide policy responses to enhance the financial protection of Georgians. These insights could also be relevant to other LMICs.

## Background

Since Georgia’s independence from the Soviet Union in 1991, it has experienced significant economic transformations, leading to economic growth and eventually becoming an upper-middle-income country. As of 2023, Georgia’s GDP per capita stood at $8,120 in current prices (5). Despite notable economic progress, poverty remains a significant concern, with nearly 20% of the population living below the national poverty line. This is compounded by income inequality, reflected in a Gini coefficient of 0.366 in 2023 (6). Progress has been made in healthcare, as indicated by Georgia’s Universal Health Coverage (UHC) service coverage index, which stood at 68 in 2021 (up from 47 in 2000), reflecting still moderate access to essential health services (7). Despite the expansion of service coverage index, households still bear a significant financial burden, as prevalence of catastrophic health expenditure due to out-of-pocket (OOP) reached 17.4 % in 2018 (8).

The epidemiological landscape of Georgia highlights the predominance of non-communicable diseases (NCDs), a trend closely linked with an aging population, accompanied with rising life expectancy (9). In 2021, cardiovascular diseases and cancer accounted for a combined 49.2% of all deaths, with the prevalence of cardiovascular diseases reaching 10,609 cases per 100,000 people and 281 per 100,000 for cancer (10). Following independence, Georgia’s health system has undergone a major transformation from the Semashko model. The system has become decentralized and was privatized through reforms carried out between 2007 and 2012 (11). In 2005, in order to reduce poverty, the government developed and implemented a proxy means-testing system that enabled the identification of low-income households with a high degree of accuracy. The administrative system than facilitated the distribution of cash transfers to low-income households and delivering healthcare benefits of the Medical Insurance Program (MIP) for poor (12). After being piloted in two regions in 2007, the MIP program was gradually expanded nationwide in 2008. By the end of 2010, nearly 21% of population were enrolled in the program, which was then further expanded to other groups of society, such as pensioners, children under five, students, teachers, military and police personnel (13,14).

In 2013, Georgia launched the Universal Health Care Program (UHCP), expanding publicly financed coverage to almost entire population. Supported by a significant increase in public health spending, these initiatives reduced financial barriers to healthcare access and increased service utilization, particularly among those who previously lacked coverage (11). Under the UHCP, inpatient and emergency care were prioritized over primary care, and coverage for outpatient medications remained limited. While the UHCP aimed to promote equity by offering greater financial benefits (no-copayment) to vulnerable groups, the complex provider reimbursement scheme enabled healthcare providers to charge patients above the UHCP tariffs (balance billing), leading to additional OOP payments (8). In 2017, the UHCP benefits were removed for the wealthiest 1% of households, and a drug reimbursement plan for non-communicable diseases was introduced, providing subsidized outpatient medications for patients with select chronic conditions—hypertension, COPD, type 2 diabetes, and thyroid disorders. Nonetheless, the financially disadvantaged continued to face OOP charges for medications (4). In 2019, the drug reimbursement plan was expanded to cover Parkinson’s disease and epilepsy. In recent years, further measures were introduced to lower medication prices, such as parallel drug imports with simplified national authorization in 2022 and a external reference pricing scheme for pharmaceuticals in 2023 (15). By 2024, user co-payments under the drug reimbursement plan were eliminated for the socio-economically disadvantaged, covering 100% of cost of a limited list of drugs (16). Furthermore, the list of pharmaceuticals subject to reference pricing has been extended to 7,100 positions and UHCP now includes coverage for a specified list of drugs used for cancer care in both inpatient and outpatient settings without annual financial limits (17). Despite the efforts, the share of OOP payments in current health expenditure in Georgia has remained relatively high over the years.

## Methodology

After securing ethics approval, the analysis used pooled data from the 2009-2023 Household Income and Expenditure Surveys (HIES) conducted by GeoStat, the Georgian Statistics Office. These surveys collect household socio-economic and consumption data using a two-stage stratified cluster sampling design, with census enumeration areas as primary and households as secondary sampling units. The sample is stratified into 21 strata based on region and urban-rural location. Around 1,440 households are randomly selected for monthly interviews. Households are interviewed twice across two quarters of a year, with follow-up interviews in the same quarters the following year. After four survey rounds, households (about 1/12^th^ of the overall panel) are replaced by randomly selected new ones from the same cluster to maintain the panel structure (18). On average, GeoStat data has 13,200 observations annually, except in 2009 and 2010 (see Table 1 for details).

**Table 1.** Characteristics of the study population, Georgia HIES 2009-2023.

| **Factors** | **Whole sample** | **Analytic sample** | **% of Analytic sample** |
| --- | --- | --- | --- |
| Total No of Records | n = 200,564 | n=198,292 |  |
| Year of HIES collection |  |  |  |
| 2009 | 22,214 | 21,859 | 11.0 |
| 2010 | 21,734 | 21,502 | 10.8 |
| 2011 | 11,170 | 11,014 | 5.6 |
| 2012 | 11,247 | 11,096 | 5.6 |
| 2013 | 11,087 | 11,004 | 5.5 |
| 2014 | 11,158 | 11,064 | 5.6 |
| 2015 | 10,992 | 10,911 | 5.5 |
| 2016 | 10,853 | 10,772 | 5.4 |
| 2017 | 11,565 | 11,531 | 5.8 |
| 2018 | 11,038 | 10,890 | 5.5 |
| 2019 | 13,863 | 13,732 | 6.9 |
| 2020 | 13,272 | 13,100 | 6.6 |
| 2021 | 13,581 | 13,391 | 6.8 |
| 2022 | 13,582 | 13,362 | 6.7 |
| 2023 | 13,208 | 13,064 | 6.6 |
| **Predisposing factors** |  |  |  |
| Household's (HH) characteristics |  |  |  |
| Sex of HH head (Male) | 132,028 | 132,020 | 66.6 |
| Higher education level of HH head (yes) | 85,383 | 85,379 | 43.1 |
| Marital status of HH head |  |  |  |
| With spouse | 123,421 | 122,293 | 61.7 |
| Without spouse | 76,984 | 75,999 | 38.3 |
| Missing | 159 |  |  |
| Age of HH head mean and (standard deviation), median | 60.9 (14.45), 61 | 61.3 (14.3), 61 |  |
| HH with at least one member < 6 years | 37,247 | 36,818 | 18.6 |
| HH with at least one member > 60 | 121,887 | 120,788 | 60.9 |
| HH size mean and (standard deviation) | 3.5 (1.9) | 3.5 (1.9) |  |
| **Enabling factors** |  |  |  |
| Per capita expenditure quintile |  |  |  |
| poorest | 44,690 | 44,331 | 22.4 |
| 2nd | 41,819 | 41,500 | 20.9 |
| 3rd | 39,954 | 39,623 | 20.0 |
| 4th | 38,548 | 38,171 | 19.2 |
| richest | 35,553 | 34,667 | 17.5 |

| Factors | Whole sample | Analytic sample | % of Analytic sample |
| --- | --- | --- | --- |
| Region of residence |  |  |  |
| Tbilisi | 29,619 | 29,399 | 14.8 |
| East | 71,601 | 70,865 | 35.7 |
| West | 99,344 | 98,028 | 49.4 |
| Residence (urban) | 82,753 | 81,959 | 41.3 |
| **Need factors** |  |  |  |
| HH with at least one member with disability | 22,886 | 22,464 | 11.3 |
| HH with at least one chronically ill member | 112,896 | 111,327 | 56.1 |
| **Expenditure on health care services** |  |  |  |
| Households with health Expenditure on: |  |  |  |
| Drugs | 140,274 | 138,769 | 70.0 |
| Out-patient treatment | 29,390 | 29,025 | 14.6 |
| In-patient treatment | 5,724 | 5,635 | 2.8 |
| Diagnostic services | 8,541 | 8,443 | 4.3 |
| Other goods and equipment | 13,116 | 12,929 | 6.5 |

GeoStat publishes anonymized micro datasets and survey documentation on its website (19). The household-level file includes variables on details of the monthly consumption using the Classification of Individual Consumption According to Purpose (20), income, and related individual and household characteristics. The individual member-level file provides demographic and socio-economic variables such as age, sex, education, and other characteristics. For the analysis, annual survey data from 2009 to 2023 were merged into a single database. For regression analysis characteristics of individual members were aggregated at the household level (our unit of analysis), either as household head attributes or relevant traits of at least one member (see Table 1 for details).

As an outcome variable, we used impoverishing health expenditures as a more nuanced concept than catastrophic expenditures and are more directly linked with poverty and inequality. Impoverishment reflects how health spending can push individuals below the poverty line or drive those who are already poor deeper into poverty (21,22). The WHO distinguishes between impoverishing health expenditures below absolute and relative poverty lines. The relative poverty line, suggested by the WHO Regional Office for Europe, accounts for basic needs like food, housing, and utilities (23), making it a more relevant measure for the European region and this research. Both impoverishing health expenditures (IHE) and further impoverishing health expenditures were combined and named as IHE and analyzed as dichotomous variable to account for both impoverishing dimensions.

Independent variables were selected based on Andersen’s model of health services utilization (24), (25) and data availability. The 200,564 households were in the pooled database, with 198,292 households (98.9%) having complete descriptors of relevant predisposing, enabling, and need factors described in Table 1 and grouped under relevant factors according to Andersen’s model. To account for the need variables and the type of health spending in the regression model, household health expenditures were converted into dummy variables separately for medicines, inpatient and outpatient treatments, and laboratory and diagnostic services and included in the model.

In this study, descriptive analysis was performed using Stata version 18 to summarize key variables and examine the distribution of the data. Survey weighted logistic regression analysis was performed in R (Version 2023.12.1+402) to assess the relationship between the variables. The approach incorporated the complex survey design, including stratification, clustering, and unequal selection probabilities, by incorporating design weights directly into the estimation process (26). The panel design of the survey was accounted for by including a household unique identifier. Various diagnostic checks were conducted before analysis to assess the nonlinearity between continuous independent variables and the log odds of IHE, multicollinearity, and outliers (27). Visual inspection indicated no evidence of nonlinearity. Variance Inflation Factor values for all variables were below the threshold of 5, indicating no multicollinearity. Cook’s distance analysis identified no extreme outliers (28). All test results are included in Supplement 1.

The factors associated with IHE were assessed using univariate and multivariate logistic regression analysis, reflected in Table 3. The association of IHE and explored factors were measured using odds ratios (OR) based on the univariate regression and the adjusted odds ratios (adj OR) in the multivariate logistic regression (MLR).

The analysis revealed that the HH expenditure quintile and drug expenditure act as explanatory variables of IHE, inflating the magnitude of the adj OR. Therefore, an interaction term between the expenditure quintile and drug expenditure was added to the final model to demonstrate a significant association between the factors and IHE. A forward selection of explanatory variables was made to address the bloated adj ORs, considering the HH expenditure quintile, in-patient hospitalization expenditure, and drug expenditure as dummy variables.

### Patient and public involvement

Patients and the public were not involved in the design, conduct, reporting, or dissemination plans of this research because the study is based on secondary anonymized data analysis, which is publicly available.

## Results

### Background characteristics of the sample

As noted in the methods section, our analytical sample comprised 98.9% of the cases in the pooled dataset, which did not introduce bias in our estimates. HIES data (see Table 1) revealed a comparable number of observations from 2011 to 2023 (5.4%—6.9%), except 2009-10, which had twice the sample size compared to the following years (10.8%-11.0%) due to the changes in the sample size that occurred in 2011.

Georgian household heads were mainly males (66.6%), had no higher education (56.9%), had spouses (61.7%), and were relatively older people, with half being older than 61 years. In terms of household composition, 18.6% had children under 5 years of age, 60.9% had members older than 60 years, 11.3% had at least one member with a disability, and 56.1% had at least one member with a chronic disease, indicating a higher burden of non-communicable diseases in the nation. Most likely, the latter determined the high prevalence of health expenditure, where 70.0% of households included in the analytic sample reported spending on drugs (likely for chronic conditions), followed by outpatient treatment (14.6%), and only 2.8% reported paying for inpatient care.

### Prevalence of Impoverishing Health Expenditure

The prevalence of IHE among Georgian households averaged 11.4 %, although this share fluctuated slightly from 2009 to 2023 without revealing any visible trend (Figure 1). The lowest recorded prevalence was in 2021 (9.8%), and the highest was in 2011 (12.8%).

**Figure 1.**
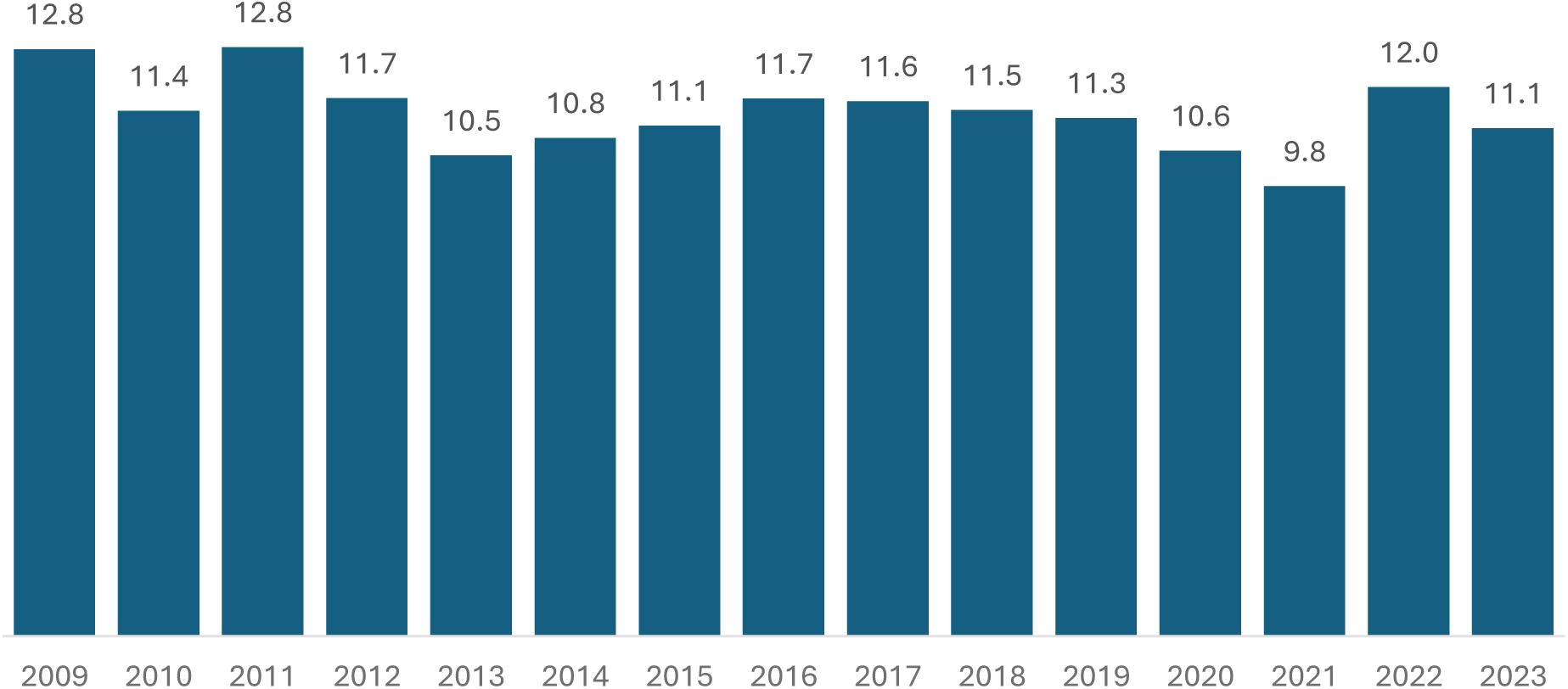
The proportion of HHs experiencing IHE over the years Georgia HIES 2009-2023

Furthermore, in Table 2, the prevalence of IHE showed significant differences by household characteristics, composition, and geographical location. The prevalence of IHE was higher among households with members under 6 years of age (13.5%), older adults (13.7%), persons with disabilities (20.3%), and those with chronic diseases (14.5%) compared to HH who reported otherwise. Additionally, households headed by females (13.6%) with no higher education (15.4%) and no spouse had a higher prevalence of IHE. Geographically, the prevalence of IHE was considerably higher in rural areas (14.2%) and in the East region (15.3%) compared to urban locations and Tbilisi.

**Table 2.** The proportion of HHs who experience IHE by factors, Georgia HIES 2009-2023.

| **Factors** |  | **Households**<br>**(analytical**<br>**sample)** | **% of IHE** |
| --- | --- | --- | --- |
| **Predisposing factors** |  |  |  |
| Sex of HH head | Male | 132,020 | 10.14 |
|  | Female | 66,272 | 13.61*** |
| HH head with higher education level | No | 112,913 | 15.35 |
|  | Yes | 85,379 | 7.33*** |
| Marital status of HH head | With spouse | 122,293 | 9.89 |
|  | Without a spouse | 75,999 | 13.73*** |
| HH with at least one member < 6 years | No | 161,474 | 10.85 |
|  | Yes | 36,818 | 13.5*** |
| HH with at least one member > 60 years | No | 77,504 | 8.13 |
|  | Yes | 120,788 | 13.69*** |
| **Enabling factors** |  |  |  |
| Per capita exp by quintile | poorest | 44,331 | 49.37 |
|  | 2nd | 41,500 | 5.95*** |
|  | 3rd | 39,623 | 0.97*** |
|  | 4th | 38,171 | 0.26*** |
|  | richest | 34,667 | 0.16*** |
| Region of residence | Tbilisi | 29,399 | 7.4 |
|  | East | 70,865 | 15.31*** |
|  | West | 98,028 | 11.58*** |
| Residence | rural | 116,333 | 14.24 |
|  | urban | 81,959 | 9.42*** |
| **Need factors** |  |  |  |
| HH with at least one member with disability | No | 175,828 | 10.35 |
|  | Yes | 22,464 | 20.27*** |
| HH with at least one chronically ill member | No | 86,965 | 7.22 |
|  | Yes | 111,327 | 14.51*** |
| **Expenditure on different healthcare services** |  |  |  |
| Expenditure on drugs | No | 59,523 | 1.56 |
|  | Yes | 138,769 | 15.6*** |
| Expenditure on out-patient treatment | No | 169,267 | 11.43 |
|  | Yes | 29,025 | 11*** |
| Expenditure on in-patient treatment | No | 192,657 | 11.45 |
|  | Yes | 5,635 | 8.93*** |
| Expenditure on diagnostic services | No | 189,849 | 11.51 |
|  | Yes | 8,443 | 8.71*** |
| Expenditure on other goods and equipment | No | 185,363 | 11.58 |
|  | Yes | 12,929 | 8.65*** |

Significance with previous row \* $<0.1$ \*\* $<0.05$ \*\*\* $<0.01$

**Table 3.**
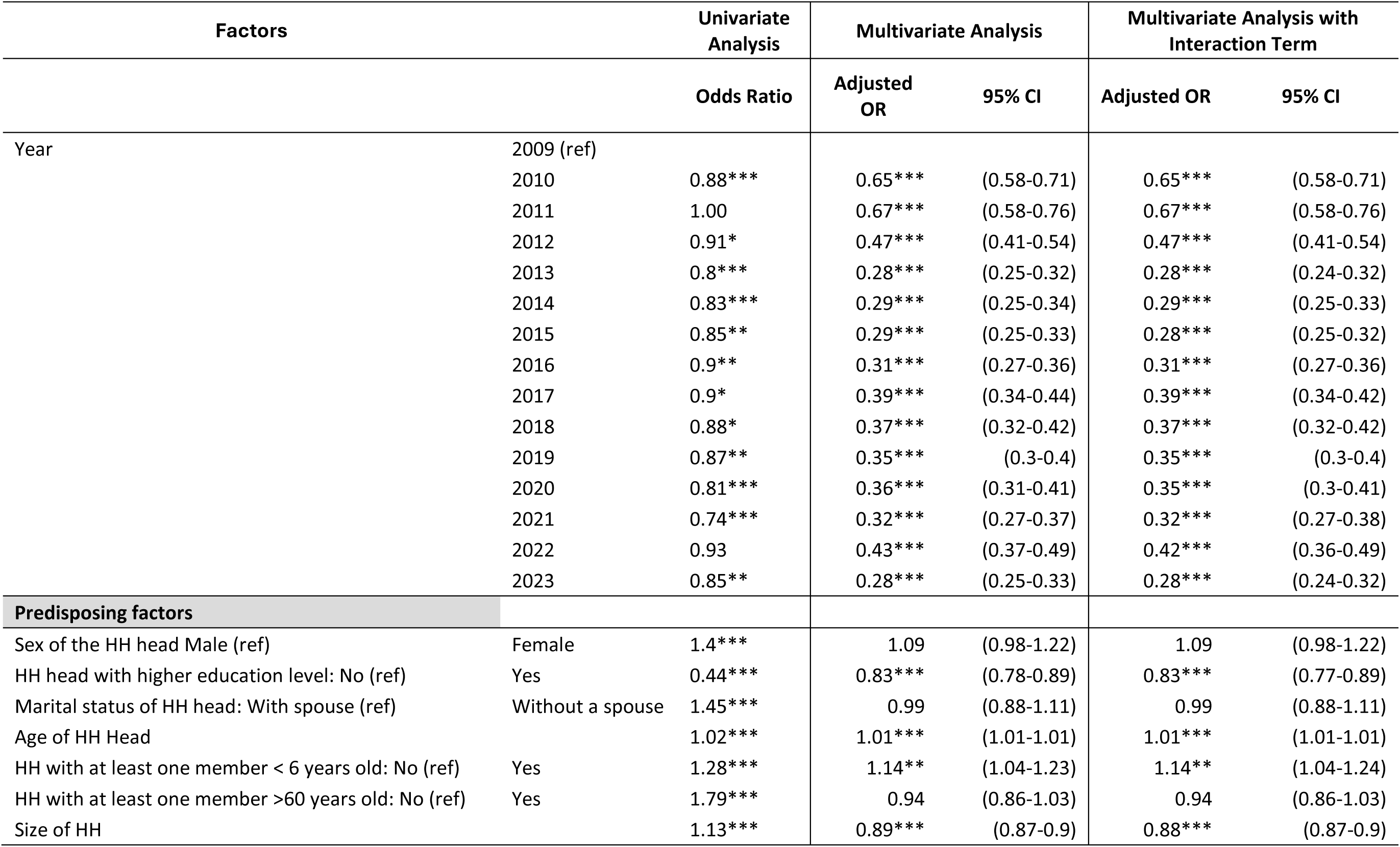
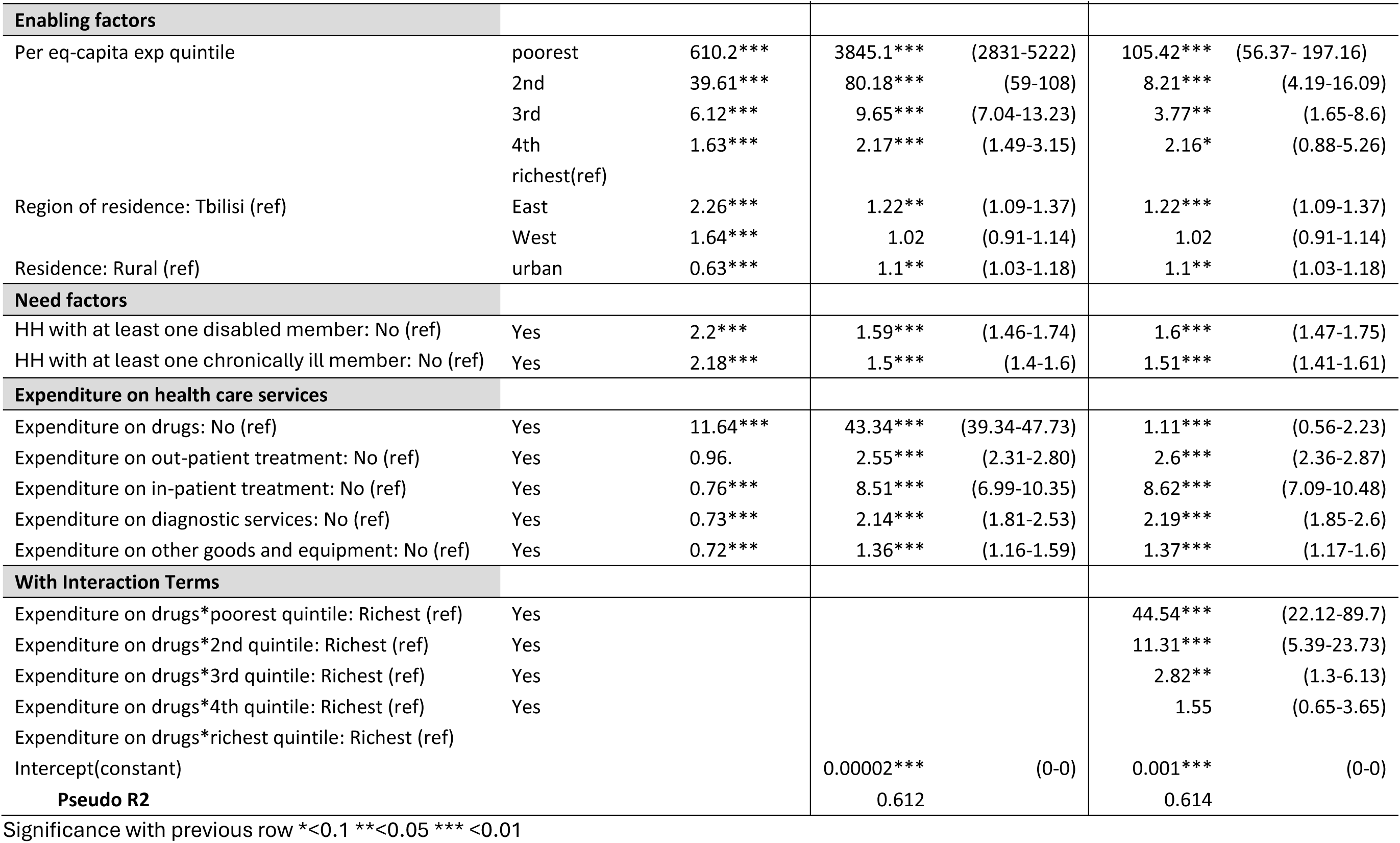
Logistic regression analysis of factors associated with IHE, Georgia HIES 2009-2023.

Disparities in the prevalence of IHE were most striking across HH’s economic status. It was alarmingly high among the poorest quintile, at 49.4%, compared to the richest, with just 0.16%. This observed difference highlights the financial strain the poorest HHs face when spending on health services.

The prevalence of IHE by specific health expenditure also exhibited significant and surprising differences. The disparity in IHE prevalence was notably high among individuals who purchased drugs (Yes: 15.6% vs. No: 1.6%). Interestingly, the incidence of IHE among households with no out-of-pocket expenditure on inpatient, outpatient, or diagnostic services surpassed those who reported spending on these services. Hence, the findings imply that medication expenses contributed more to household financial hardship than other expenditure types.

### Factors associated with Impoverishing Health Expenditure

The univariate logistic regression showed that all identified factors were significantly associated with IHE, except for households with outpatient treatment expenditures (see Table 3). However, with univariate analysis alone, we did not draw definitive conclusions. Therefore, multivariate regression testing of these factors was necessary. In the multivariate regression analysis, the association of these factors with IHE underwent significant changes. The most noticeable differences between the OR and the adj ORs were for the poorest household expenditure quintile, where the adj. OR disproportionally increased for these households. This was followed by drug and inpatient expenditure. The results of the MLR revealed a potential mediating effect involving household expenditure quintile, inpatient hospitalization expenditure, and drug expenditure in their association with IHE, given the inflated magnitudes of the adjusted odds ratios. Our multivariate analysis confirmed several factors from the literature that influence household financial hardship levels (29). These include the low educational attainment of a household head and his/her age, households with young children under six, HHs with senior members over 60, or those with a disabled or chronically ill member. The impact of these factors on the likelihood of IHE was generally modest, except the presence of a member with disability, which increased the odds by 59% (OR-1.59 (95% CI: 1.46-1.74)), and a HH member with chronic condition raised them by 50% (OR-1.50 (95% CI:1.40-1.60)).

Next, our analysis has revealed significant changes along the time trend for odds of impoverishment due to health spending. While the proportion of impoverished households remained relatively stable over the study period (See (Figure 1), our multivariate analysis has shown a noticeable decrease in the odds of impoverishment, holding all other factors constant, between 2009 and 2013 (see Table 3). During this period, the odds dropped to their lowest level in 2013 at OR-0.28 (95% CI: 0.25-0.32) compared to 2009. Post-2013, although the odds were consistently lower than in 2009, they did not reveal further decline. On average, the adjusted odds ratio from 2014 to 2023 stood at around 0.34.

Furthermore, when controlling for all other factors, the poorest 20% of households were significantly more vulnerable, with odds of impoverishment 3,845 times higher (95% CI: 2831-5222) than the wealthiest 20%. In contrast, the second quintile showed dramatically – forty-eight times lower odds – 80.18 (95% CI: 59-108), indicating profound disparities in financial risk due to health expenses between the poorest and the second quintile.

Furthermore, the analysis of health expenditure types reveals that the cost of purchased medicines has the most impoverishing impact on households, with an adj OR of 43.34 (95% CI: 39.34-47.73), compared to the subsequent most impoverishing expenses for inpatient/hospital treatment, with an OR of 8.51 (95% CI: 6.99-10.35). For all other healthcare spending, odds ratios were around two or below.

To conclude, our analysis of healthcare expenditures reveals a significant finding: households with chronic individuals or disabled persons and being at the bottom of the income ladder and having expenses on drugs face significantly higher odds of impoverishment compared to those paying for inpatient (or other) services or who are wealthier. This indicates that pharmaceutical costs are a primary driver of healthcare-related impoverishment, especially among the poorest, where almost half face IHE.

## Limitations

The findings presented should be interpreted with the following limitations in mind. In the logistic regression analysis, household need was represented by dichotomous variables based on reported expenditures for specific services (e.g., inpatient, outpatient, medicine). This was necessary because the questionnaire did not include a question about the actual need for healthcare. As a result, we only capture households that both needed and accessed care and paid for services. Our analysis did not account for households that may need care but could not access or pay for it. However, this does not bias our results since the focus is on the impoverishment caused by healthcare payments, which cannot be assessed without such payments.

Additionally, our calculation of impoverishing healthcare expenditure only considers the burden of direct treatment costs. Indirect costs, such as transportation to healthcare facilities or income loss due to treatment, were not accounted for. While including these costs would likely have increased the overall level of impoverishment, it is unlikely to have significantly altered the regression results.

## Discussion

WHO advocates for UHC, which Georgia has implemented since 2013. With a broadened benefit package, 90% of the population was covered. However, this expansion did not improve financial protection as measured by the IHEs, particularly for the poor, a challenge also observed in other countries (30).

Several factors could be at play. Firstly, the expansion of UHC in Georgia primarily focused on increasing inpatient benefits compared to other services. The highest budget allocations were made for inpatient services, which grew fast over the past decade (31). During 2009-2023, overall public spending on health grew faster from 0.35 billion Gel to 1.1 billion (and increased more than 3 times), which was accompanied by a reduction in the share of out-of-pocket payments in the current health expenditure from 68.91% in 2009 to 40.45% in 2022 (31,32). Consequently, compared to a previous study from 2009 (33), the population in 2023 seems better protected against hospitalization expenses, as the odds of catastrophic payments and consequent impoverishment appear to have declined significantly. Most likely, this highlights a positive outcome of UHC implementation, which helped safeguard the patients from the impoverishing effects of inpatient care costs, while improving access to inpatient services for those in need (4). However, offering outpatient drug benefits has been postponed for some years since 2013 (34). The delayed inclusion of pharmaceutical coverage most likely harmed the levels of impoverishment, which were not reduced as expected.

As noted earlier, the reforms in pharmaceutical coverage took some years and only started in 2017, albeit with poorly defined benefits, small budgetary allocation, and weak implementation arrangements that led to low program uptake by beneficiaries (35). Impactful reforms in pharmaceutical benefits only picked up in 2020 when the package began covering treatments for four non-communicable conditions (NCD), expanded covered population groups, and streamlined implementation arrangements (36). This led to a greater uptake of NCD medicines, which are undoubtedly beneficial in reducing outpatient drug costs for the population.

Nonetheless, state-funded pharmaceuticals are still a small share (about 17% in 2023), of total pharmaceutical expenditures, with the population bearing 83% of these costs. Conversely, the state budget is the predominant funding source for inpatient services—81% in 2023, and the population only pays 19% (19,37). Although Georgia is a middle-income country, the expectation that the government’s fiscal capacity will allow a significant shift in this public-private balance for pharmaceutical payments in the near future is less likely without a major change in the government’s overall budgetary priorities.

The significant reduction in the odds of impoverishment observed during 2009-2013, followed by no progress after that, suggests that early government interventions—likely through targeted, mostly emergency outpatient and planned and emergency inpatient benefits for the poorest and pensioners — had a higher protective financial effect. Particularly when the initial barriers to coverage were removed by 2012, and most of the intended beneficiaries were able to benefit (38). However, the pace of reduction in odds was not sustained by later reforms after the implementation of the UHCP. The government shifted towards universal coverage by extending outpatient and inpatient benefits to include other, relatively better-off population groups, who may have experienced less impoverishment as a result of the expenditures on these services, without expanding coverage for another major determinant of IHE -outpatient drug expenditures - for the poor and better-off alike. Additionally, this transition from the targeted to the universal approach in a context of limited funding may have disproportionately benefited the better-off at the expense of more poor or vulnerable, who received less targeted support.

Although there is considerable controversy in published literature around universal arrangements versus targeting public subsidies to poor or vulnerable groups, we argue that going forward in Georgia and improving coverage for targeted population groups with greater needs, including people with chronic conditions from low-income households, could help reduce impoverishment resulting from health spending. While solid arguments exist in favor of a well-funded, publicly led, and universal approach to coverage (39,40), Georgia’s case reveals that UHCP has delivered on its objectives for inpatient services but not for pharmaceuticals because the universal approach has failed to provide adequate financial protection for the lowest income groups. Given current resource limitations, “progressive universalism,” coined by Gwatkin and Ergo, (41) looks more appropriate for Georgia’s context. It seems necessary to target the expansion of the state-funded pharmaceutical benefits to those who face the highest financial risk (i.e., the poorest 20% of the population) who may not have the greatest need compared to the pensioners, but even a small amount of payment for prescribed medicines could have an impoverishing effect and push almost half of these households into poverty or further impoverish them. Therefore, our findings suggest that the government should focus on and significantly expand pharmaceutical subsidies for the poorest as the initial step before considering other population groups for expansion. This suggestion builds more on recommended solutions to target resources to the poorest (42,43), noting limitations that there is little evidence that targeting is cost-effective relative to broader approaches to service provision (44,45). However, as mentioned earlier, Georgia has an administrative mechanism to identify the poor and deliver state assistance, and implementing this suggestion seems rather feasible in a given context.

In conclusion, achieving the Sustainable Development Goals of Universal Health Coverage requires more granular analysis and adaptation to each country’s context based on this evidence. Simply increasing coverage is insufficient to meet UHC goals without closely examining the local circumstances, determining which benefits should be prioritized, and identifying the population groups that would benefit most. No country has the resources to meet all health needs, so trade-offs are inevitable (46). Future healthcare reforms in Georgia and elsewhere should focus on expanding coverage and improving equity, with an emphasis on identifying the benefits that could provide the greater financial protection for the poor and other vulnerable groups. This approach could help ensure equity while progressing toward UHC goals.

## Concluding Remarks

While our study provides quantitative insights into impoverishing drug expenditure in Georgia, to better shape the policies, a more in-depth qualitative assessment of pharmaceutical benefits offered by the state would be necessary. Specifically, it would be important to understand why the efforts of 2017-2019 have not delivered on expectations and what has changed since 2020 that facilitated increased uptake of pharmaceutical benefits by the public; and why even recent policy and implementation changes since 2020 are not able to reduce odds of impoverishment and what could be changed on a policy or implementation level to achieve better outcomes, especially for the poor.

## Data Availability

All the data used is available in the public domain: https://www.geostat.ge/en/modules/categories/128/databases-of-2009-2016-integrated-household-survey-and-2017-households-income-and-expenditure-survey.

## Funding Statement

This study was funded by the Shota Rustaveli National Science Foundation of Georgia under grant number FR-22-7764. The sponsors were not involved in the study’s design, data collection, analysis, interpretation of results, or the decision to publish.

## Author contribution statement

Contributions: T.G., A.Z. and G.G. conceptualized the paper, selected a theoretical framework and methodology for the analysis. T.G. collaborated with J.M.C. on the descriptive and inferential analysis and implemented selected methodology to answer the research objectives. J.M.C. performed the descriptive analysis and ran the initial regression analyses and diagnostic tests. T.G. focused on the final logistic regression and model testing. G.G., T.G and A.Z reviewed and verified the analytical methods and obtained results.

All authors contributed to the manuscript production: J.M.C. contributed to the descriptive analysis section. T.G., A.Z. and G.G. authored the introduction and background sections and supported the writing of logistic regression results. G.G. and A.Z contributed to interpreting the results. All authors provided critical input, shaping the research, analysis, and manuscript production.

## Competing interest statement

All authors declare that they have no competing interests.

## Ethics approval

Although this was secondary analysis of the publicly available anonymized datasets, the research team did obtain the ethical approval from the Health Research Union’s ethical committee (protocol #2024-01, approved 04/03/2024).

## Data sharing statement

The data used in this research is available from the GeoStat website: https://www.geostat.ge/en/modules/categories/128/databases-of-2009-2016-integrated-household-survey-and-2017-households-income-and-expenditure-survey.

## Notes

### Competing Interest Statement

The authors have declared no competing interest.

### Author Declarations

The study used ONLY openly available human data that were originally located at:https://www.geostat.ge/en/modules/categories/128/databases-of-2009-2016-integrated-household-survey-and-2017-households-income-and-expenditure-survey.

